# Evaluating the COVID-19 responses of Belgium, Denmark, Germany, the Netherlands, Sweden and the United Kingdom, February-June 2020: A counterfactual modelling study

**DOI:** 10.1101/2024.07.05.24309992

**Authors:** Pieter T. de Boer, Fuminari Miura, Giske R. Lagerweij, Jacco Wallinga

**Author notes:** Corresponding author: Pieter T. de Boer, dr., Postbus 1 (box 75), 3720 BA, Bilthoven, the Netherlands.

## Abstract

**Introduction:** Differences in responses to the COVID-19 pandemic among Northwestern European countries have generated extensive discussion. We explore how the impact of the first pandemic wave might have differed, had Belgium, Denmark, Germany, the Netherlands, Sweden and the United Kingdom adopted responses from the other countries, or had it delayed its own response.

**Methods:** The time-varying reproduction number R_t_ for each country was estimated using time-series of laboratory-confirmed COVID-19 deaths. Counterfactual assessment of the impact of responses was conducted by interchanging the reduction in reproduction number by calendar date between countries from March 13^th^ to July 1^st^, 2020. The impact of a delayed response was evaluated by lagging the time-series of the reproduction number with one day or three days.

**Results:** The cumulative number of COVID-19 deaths for any of the six countries would have differed substantially, had the response of another country been adopted on the respective calendar date. The order, from the lowest to the highest expected mortality rate, was obtained with the responses of the Netherlands, Belgium, Denmark, the United Kingdom, Germany, and Sweden, with a seven- to twelve-fold difference between the lowest and highest outcome. For the Netherlands, delaying its response by three days resulted in a doubling of the cumulative COVID-19 mortality rate.

**Conclusion:** During the fast-growing first COVID-19 wave, small differences in initial epidemiological situations between countries, together with small differences in the timing and effectiveness of adopting COVID-19 response from neighboring countries, result in large variations in mortality rates.

## Introduction

The first pandemic wave of coronavirus disease 2019 (COVID-19) cases from February to June 2020 led to varying health impacts across Northwestern European countries. For instance, Belgium and the United Kingdom (UK) experienced approximately eight times more confirmed COVID-19 deaths during this period compared to Germany and Denmark [1]. The difference in COVID-19 response between those countries could have played a crucial role in these variations [2, 3]. All Northwestern European countries implemented sets of non-pharmaceutical interventions (NPIs), although with varying timing of introduction and stringency [4]. These NPIs ranged from restrictions on mass gatherings and social contact, along with the closure of schools, bars, and restaurants, to less strict, voluntary measures while keeping schools, bars, and restaurants open with certain restrictions.

For each country, debates emerged on what the outcome would have been, had a different response been used. However, to quantify the impact of diverse responses in a country, one needs to rely on a modelling approach, and one needs to determine alternative, counterfactual strategies. An infinite number of counterfactual strategies are possible involving different combinations of NPIs, varying timings of implementation and relaxation, and diverse levels of compliance. In this context, instead of testing every hypothetical combination, we choose to compare strategies that were actually implemented in a selection of countries of interest. This methodological choice allows for more data-driven assessment of the effectiveness of NPIs, measured as a reduction in reproduction number R_t_, as outlined by recent studies [5, 6].

In this study, we explore the impact of counterfactual responses as implemented in Belgium, Denmark, Germany, the Netherlands, the UK, and Sweden on COVID-19 mortality during the first wave of the pandemic. These countries were chosen for their similar socio-economic characteristics and minor discrepancies in factors such as the timing of SARS-CoV-2 introduction, while showing some variation in the timing of introduction and strictness of the response measures. The study includes all 30 comparative analyses between all 6 countries; for the sake of presentation, we will first focus on the outcome for the Netherlands and then highlight differences in outcomes for other countries. To distinguish the impact of difference in response timing from differences in selected NPIs, we conduct an additional analysis examining the impact of delaying the implementation of the response.

## Methods

### Analysis framework

We used the counterfactual modeling framework developed by Mishra et al. [5] for each of the six countries in the period February to June 2020, had it used another countries response. In this framework, the time-varying reproduction number R_t_ was first estimated using time series of daily laboratory-confirmed COVID-19 deaths by date of death. We chose confirmed deaths as an outcome, as it is available for all selected countries and it is less influenced by differences in testing policies compared to confirmed COVID-19 cases. In the counterfactual analysis, the relative reduction in the reproduction number (R_t_ with control measures relative to the R_t_ without control measures) is taken from one country, and applied to the other countries. Such an approach enables the transfer of both the timing and the magnitude of reduction of the transmission intensity (i.e., the response effectiveness), while maintaining the country-specific features upon which R_t_ without control measures is based, such as population density and international connectivity.

### Data

Time series data on daily deaths by date of death for Belgium, Denmark, Sweden and the UK up to 1 July, 2020, were obtained from a public source [7] or from Mishra et al. [5]. For the Netherlands, such data were extracted from the OSIRIS database, the national registry for laboratory-confirmed COVID-19 cases of the Dutch National Institute for Public Health and the Environment, with deaths with missing date of death omitted. For Germany, data was received from the Robert Koch Institute (personal communication, Matthias an der Heiden, 1 December 2022). Consistent with observed serial interval for SARS-CoV2 transmission in the Netherlands, we used a generation time that followed a gamma distribution with a mean of 4 days and a standard deviation (SD) of 2 days [8]. The mean of the infection-to-death delay distribution was assumed to be the same between countries, using the sum of the infection to onset duration (approximate mean of 5.2 days with SD of 2.2 days [9]) and the onset to death duration as estimated for England (approximate mean of 15.1 days with SD of 12.6 days [5]). Other parameter values that were used in the analysis can be found in [5].

### Estimation of the reproduction number

The reproduction number R_t_ for each country was estimated by fitting a semi-mechanistic transmission model [5] to the time series of daily deaths. R_t_ is defined here as the instantaneous reproduction number [10, 11], calculated as the number of new individuals infected by a single infectious person who is infected at time t. R_t_ without control measures was estimated by fitting the model to the first week of time-series of deaths after a country had observed a total of 10 cumulative death cases. This ensures that the deaths were not caused by imported infections but also were not affected by control measures, given the delay between infection and death.

### Counterfactual assessment

For each country, a growing epidemic was simulated until March 13, 2020, maintaining a value for the reproduction number as observed in the specific country without control measures. This date marks the point at which countries in Northwestern Europe started taking stringent control measures. Subsequently, from March 13 to July 1, 2020, we substituted the relative reduction in the reproduction number of the one country (‘recipient country, e.g. the Netherlands) by that of another country (‘donor’ country, e.g. Belgium) on the corresponding calendar day. We repeat this approach for each country, and substituted the relative reduction in the reproduction number as observed for other countries, and assessing the consequences in terms of mortality.

For the Netherlands, we also assessed the impact of a delayed response, by shifting the observed time-series of deaths to one day or three days later, computed the corresponding (lagged) reproduction number R_t_, and interchanged the values of the relative reduction in the reproduction number without control measures as in the between-country comparison.

## Results

### Time course of observed COVID-19 mortality per country

The observed cumulative mortality rate of 1 per million was reached first in the Netherlands, Belgium and the UK (14 March), and later by Denmark (17 March), Sweden (18 March) and Germany (20 March) (Figure 1). Whereas the mortality rates for most countries remained close to zero after May, 2020, the rate for Sweden was relatively high. This resulted in cumulative mortality rates that were almost constant for most countries after May 2020, and for Sweden a steady increase.

**Figure 1:**
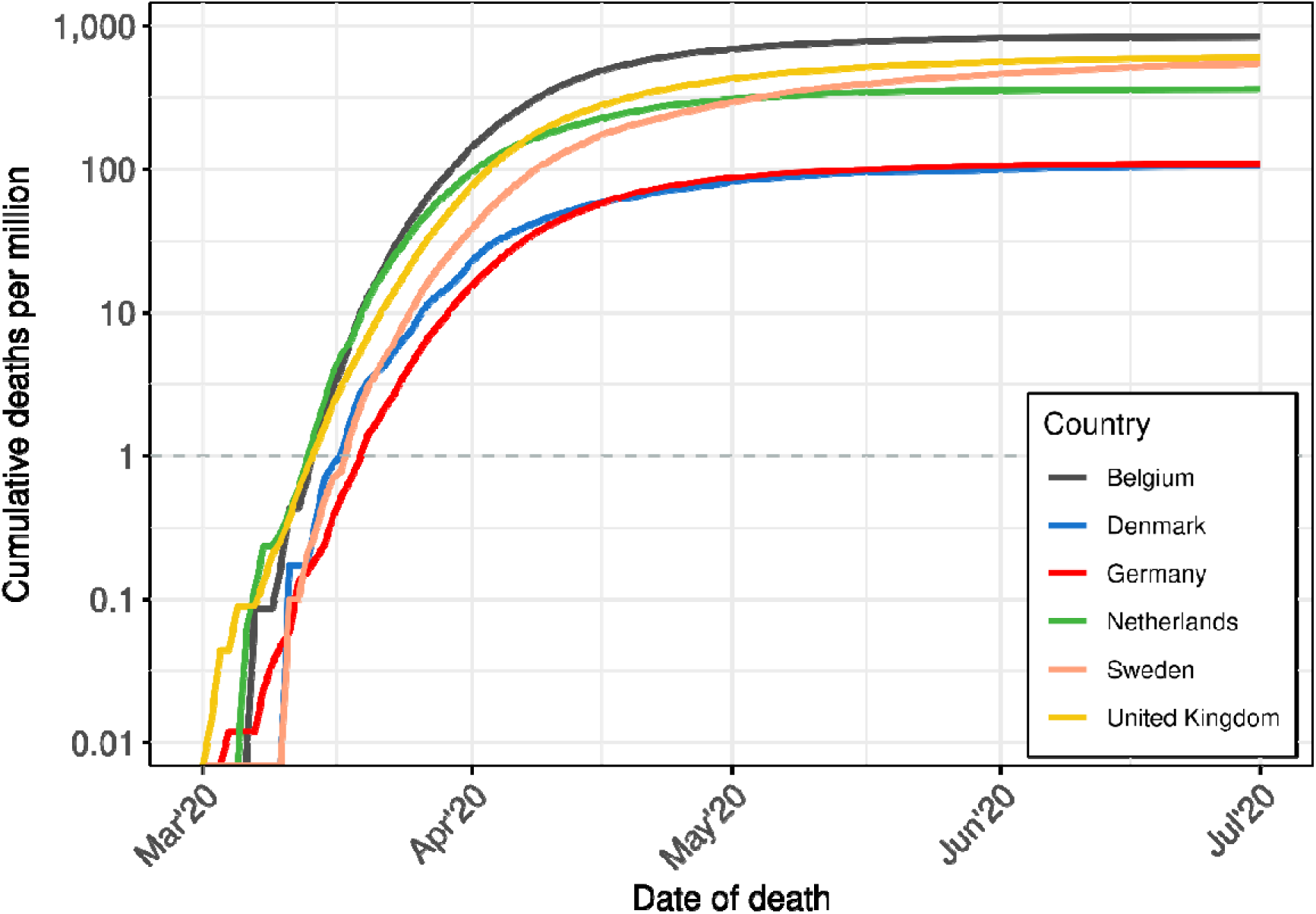
Cumulative laboratory-confirmed COVID-19 deaths per million by date of death per country, in the period March 2020, up to and including June, 2020. The grey dotted line represents the level of 1 death per million inhabitants.

### Time-varying reproduction number (R_t_) per country

The median reproduction number R_t_ without control measures for the Netherlands was estimated at 3.7 (see Supplemental Figure S1, R_t_ before March 13). With a mean generation time of 4 days, this means that the number of infections doubles approximately every 2.1 days. The other countries, in descending order for the reproduction number without control measures, are Belgium (3.7), the UK (3.6), Germany (3.5), Sweden (3.3), and Denmark (3.2).

After the introduction of control measures on March 13, the reproduction number R_t_ for the Netherlands dropped to 1.8 by the week of 13-19 March, and further to 0.9 by 20-26 March; the critical threshold of 1 was surpassed with an absolute drop of 0.9 per week (from 1.8 to 0.9). Denmark also crossed the threshold in the same week, albeit reaching a higher absolute value of the reproduction number (R_t_ ≈1) and at a slower rate (absolute weekly drop of 0.8). Belgium, Germany, the UK, and Sweden surpassed the threshold one week later, by March 27-April 2, with absolute drops in the range of 0.5 to 0.6 in that week.

The lowest value for the reproduction number R_t_ for the Netherlands was reached by April 10-16 (R_t_ of 0.6). The other countries, in increasing order of the lowest values for R_t_ were Belgium (0.5), Germany (0.6), Denmark and the UK (0.7), and Sweden (0.8) in the first half of April. The reproduction number R_t_ increased in all countries in the second half of April, Denmark experienced a relatively smaller rise. By June, Belgium and Germany had a reproduction number R_t_ around 1, while other countries saw a decline in reproduction number.

### Counterfactual assessment of country-specific response strategies

For the Netherlands, the actual decline in reproduction number R_t_ (Figure 2, blue lines) was faster from 13 March onwards compared to the decline in the reproduction number R_t_ corresponding to the counterfactual responses of the other countries in the Netherlands (Figure 2, red lines). For most of the other countries, the drop in R_t_ lagged only a few days; the decline in R_t_ corresponding to the response of Sweden, was substantially slower. The responses of Denmark and Sweden corresponded to a drop in the reproduction number R_t_ that was less far below 1 as compared to the drop in reproduction number corresponding to other countries.

**Figure 2:**
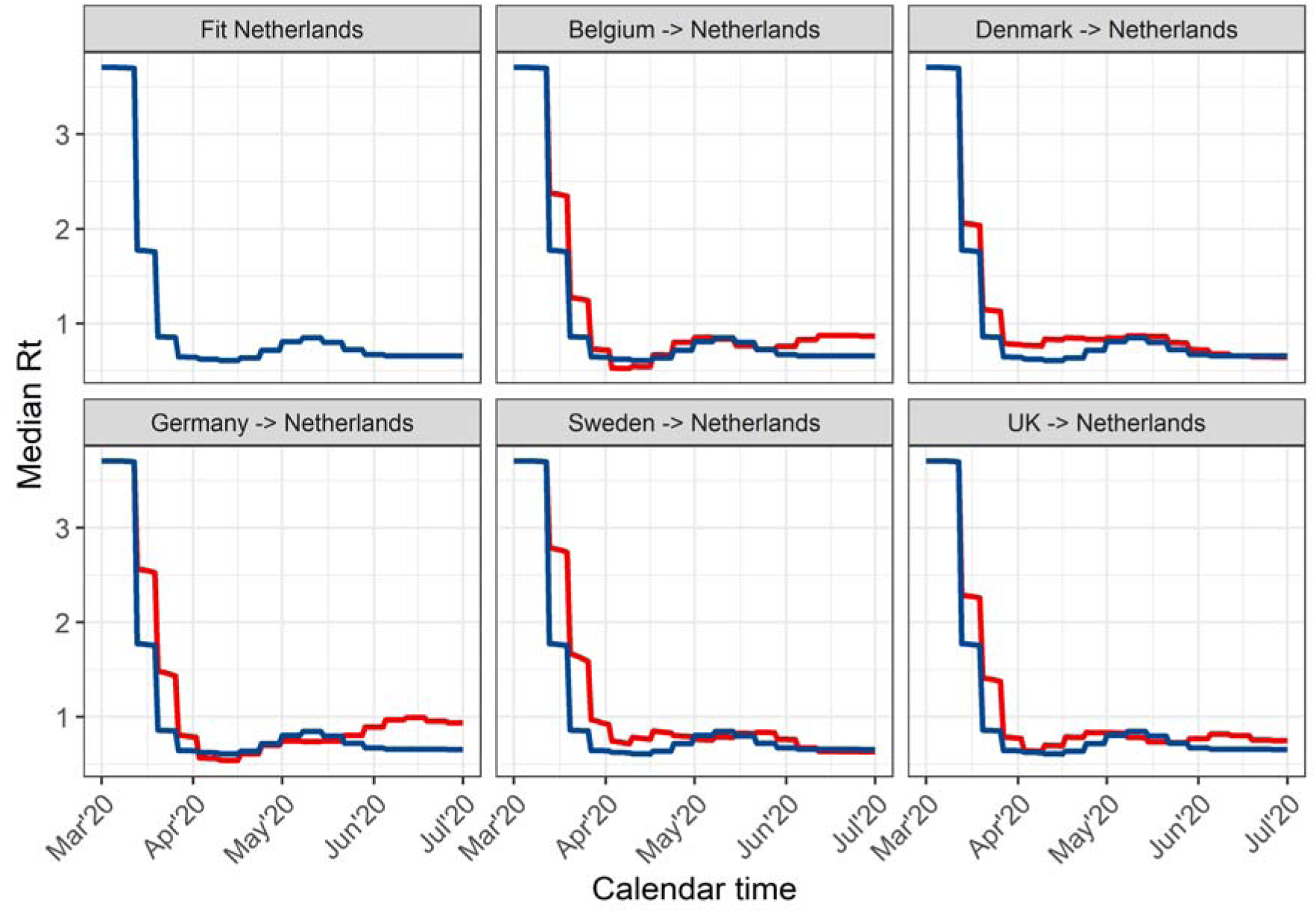
The estimated median reproduction number R_t_ for the Netherlands (blue lines) using mortality data, and the R_t_ for the different counterfactual analyses (red lines), after transferring the relative reduction in reproduction number from Belgium, Denmark, Germany, Sweden, and the United Kingdom (UK) to the Netherlands between March 13, 2020, and July 1, 2020.

The slight variations in the reproduction number R_t_ between countries have significant implications for the mortality rate. With strategies as implemented in Denmark, Belgium, Germany and the UK, the peak mortality rate in the Netherlands during the first wave would have surged from approximately 10 per million population per day to a range of 18 to 35 deaths per million population per day (Figure 3, based on median estimates). The response as implemented in Sweden increased the peak daily deaths in the Netherlands to nearly 55 per million population per day. For any of the responses implemented in the five other countries, the counterfactual cumulative deaths per million during the first wave in the Netherlands would been significantly higher than the actual mortality as observed with the actual response (Table 1). The response strategies as implemented in Belgium and Denmark resulted in a two-fold increase in cumulative deaths per million compared to the observed in the Netherlands, while the response strategies as implemented in Germany and UK led to a three-fold increase in cumulative deaths per million. The response as implemented in Sweden was even associated with a seven-fold increase in deaths per million.

**Figure 3:**
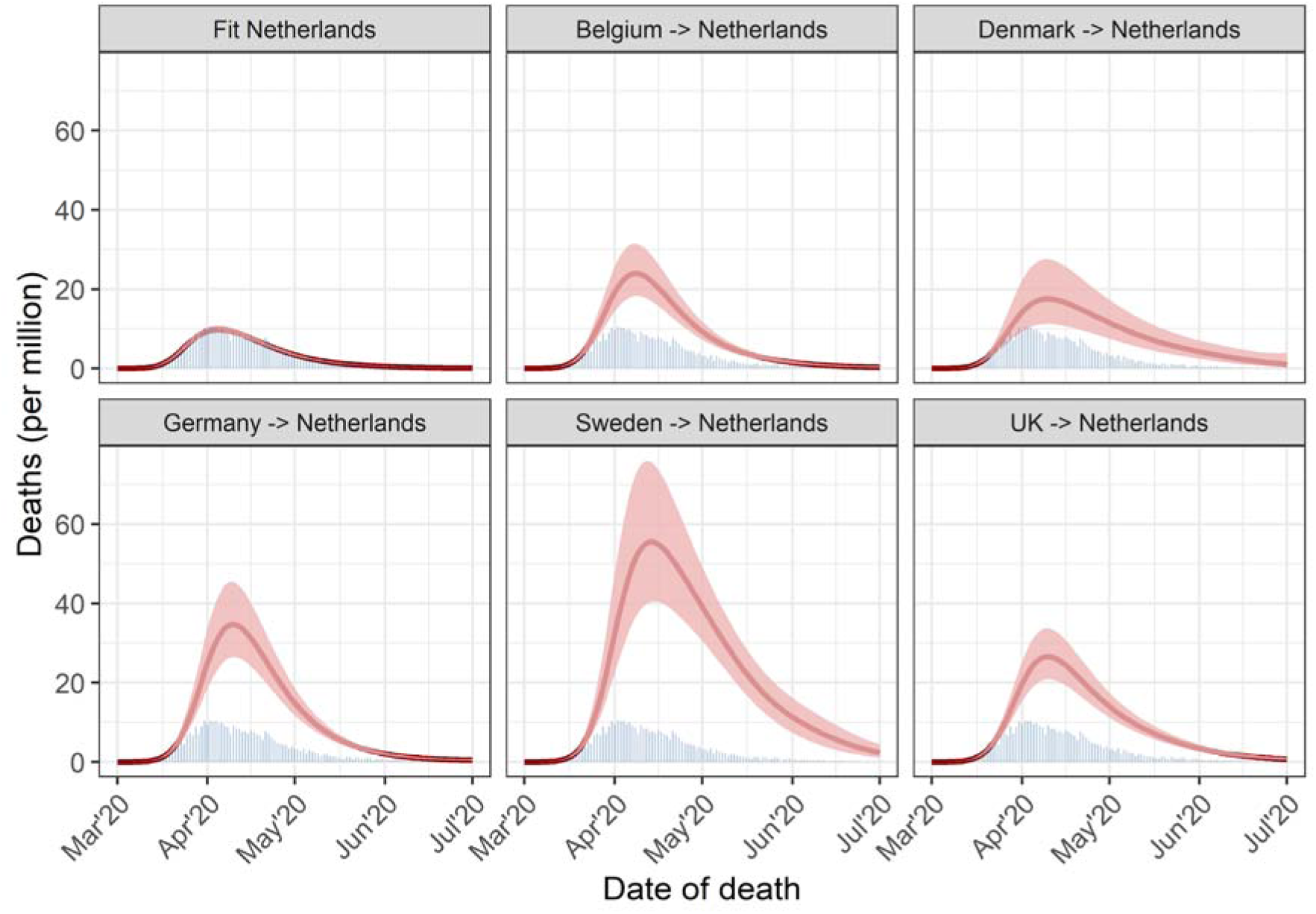
Estimated median number of daily deaths per million population with 95% credible intervals for the Netherlands, showing the fit to observed data (blue bars) and the counterfactual analyses, involving the transfer of the relative reduction in reproduction number from Belgium, Denmark, Germany, Sweden, and the United Kingdom (UK) to the Netherlands in the period March 13, 2020, to July 1, 2020.

**Table 1:**
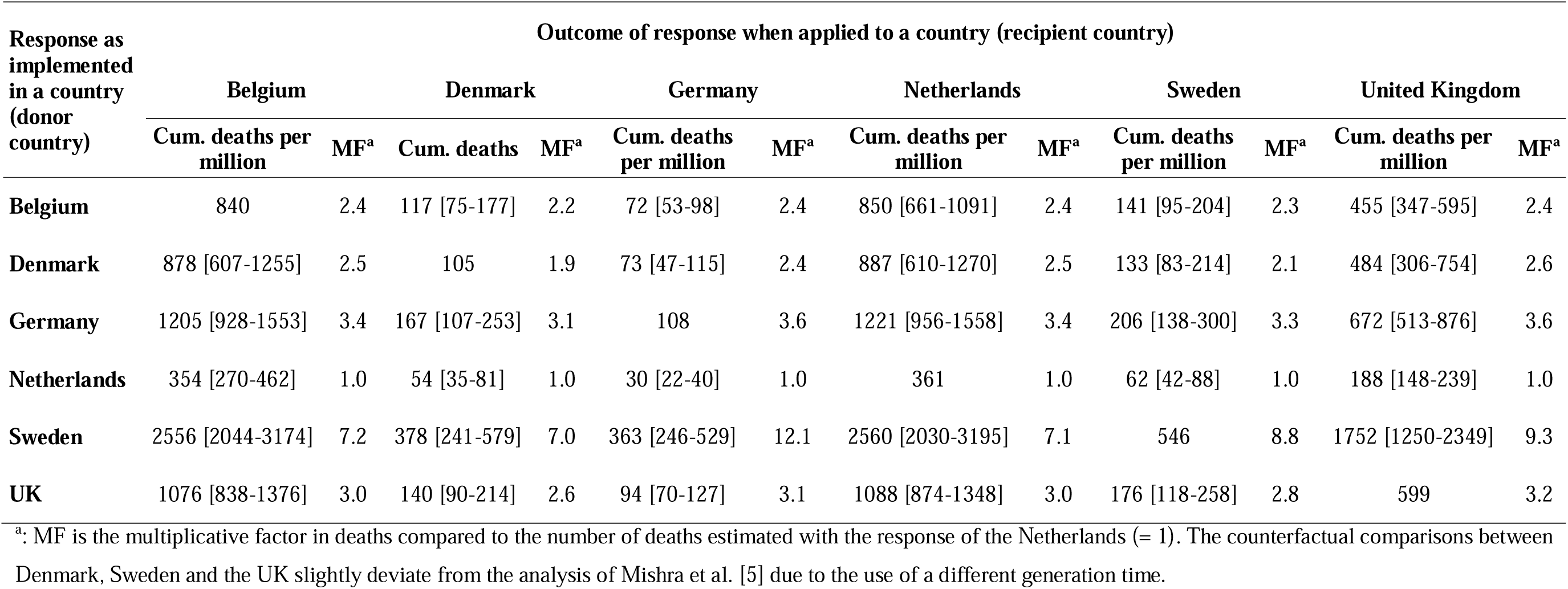
Cumulative COVID-19-attributed deaths per million inhabitants per country until 1^st^ of July 2020. Diagonal elements show observed mortality, off-diagonal elements show mortality (median with 95% credible intervals) for counterfactual strategies, using the reduction in R_t_ with control measures relative to the R_t_ without control measures. In this approach, the relative reduction in the reproduction number R_t_/R_o_ from the ‘donor’ country is transposed to the ‘recipient’ country, starting on 13^th^ March, 2020. The multiplication factors visually show per column how the mortality of the counterfactual response would have changed relative to observed mortality. A factor above 1 indicates an increase in deaths with the counterfactual response, while a factor below 1 indicates a decrease.

The order of countries’ responses with respect to the expected cumulative COVID-19 deaths was consistent when applied to the various countries; the response of the Netherlands yielded the fewest deaths per million, followed by Belgium and Denmark, then the UK and Germany, and finally Sweden (Table 1, counterfactual time series of reproduction number R_t_ and time series of mortality rates for each country shown in the Supplemental materials: Figure S2-Figure S11). In Denmark and Sweden, the response of Denmark resulted in lower mortality rate compared to the response of Belgium; for Belgium, Germany, the Netherlands and the UK, the response of Belgium resulted in a lower mortality rate compared to the response of Denmark. However, the multiplication factor for cumulative deaths per million depends not only on the response of a donor country but also on the recipient country. Whereas for the Netherlands the ratio between highest and lowest mortality amounted to a seven-fold difference, for the UK this was a ten-fold difference, and for Germany this was a twelve-fold difference.

### Counterfactual assessment of delaying the response

Delaying the response as implemented in the Netherlands by one day or three days increased the peak number of deaths in the Netherlands from approximately 10 per million population per day to 12 per million population per day and 23 per million population per day, respectively (Figure 4, see Supplemental Figure S12 for the reproduction number R_t_ profiles). The number of deaths throughout the first wave was estimated to increase by a factor 1.2 (95% CrI: 0.9-1.6) for a one-day delay, and a by factor 2.3 (95% CrI: 1.7-3.1) for a 3-day delay (Table 2).

**Figure 4:**
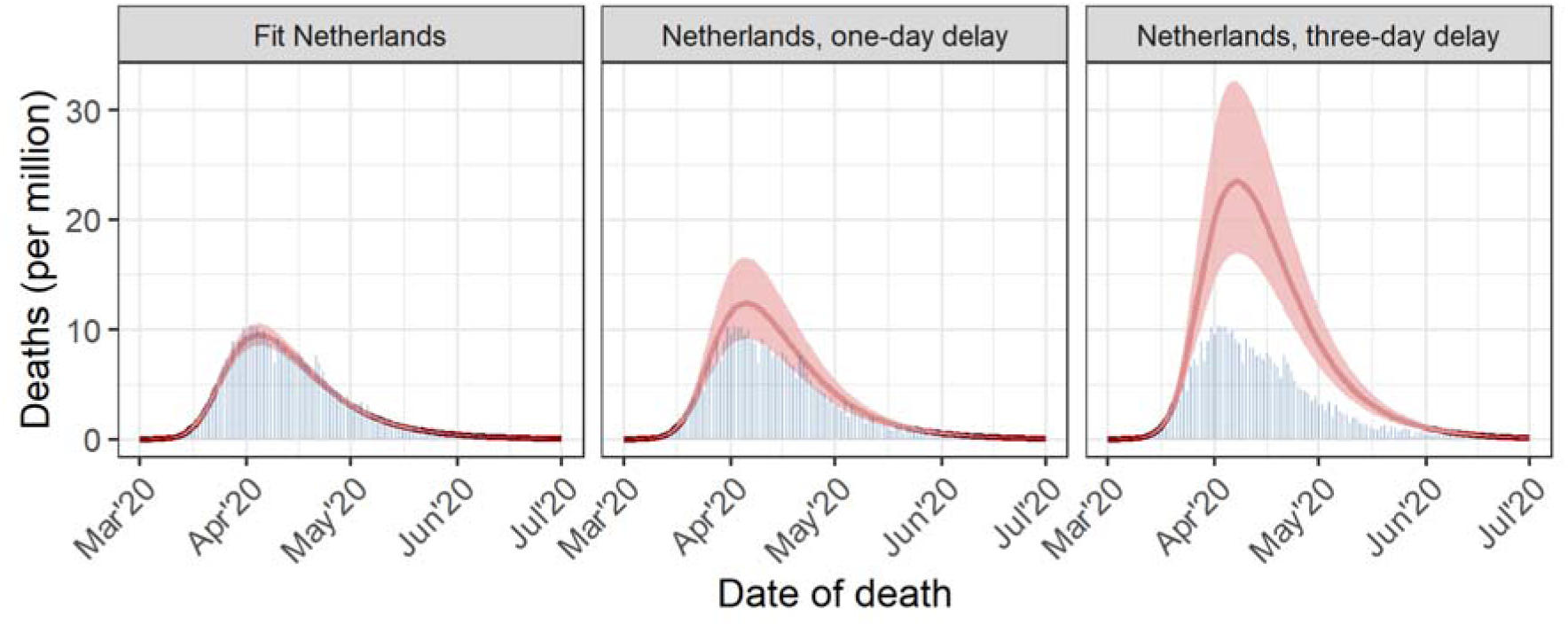
Estimated median number of daily deaths per million population with 95% credible intervals for the Netherlands, showing the fit to observed data (blue bars) and the counterfactual analyses involving response measures taken one day later or three days later in the period March 13, 2020, to July 1, 2020.

**Table 2:** Estimated relative differences in cumulative deaths per million during the first COVID-19 wave, compared to observed number of deaths, if the response of the Netherlands had been delayed by one day or delayed by three days. The analysis covers the period February to June 2020, with counterfactual strategies involving the transfer of the relative reduction in reproduction number from different countries to the Netherlands from March 13, 2020, onwards. If the multiplication factor is above 1, the number of deaths of the Netherlands would have increased with the counterfactual response strategy, and if below 1, the number of deaths would have decreased.

| **Counterfactual COVID-19 response** | **Multiplication factor of cumulative deaths,**<br>**median (95% credible interval)** |
| --- | --- |
| Actual response | Reference (=1) |
| One-day delay of response | 1.2 (0.9-1.6) |
| Three-day delay of response | 2.3 (1.7-3.1) |

## Discussion

This analysis quantifies the potential impact of different pandemic response strategies on COVID-19 mortality in Northwestern European countries during the first pandemic wave in February through June, 2020. It highlights that in the rapidly growing first COVID-19 wave LJ infection rates initially doubled every 2-3 days LJ, small differences in initial epidemiological situations between countries, together with small disparities in the timing and effectiveness of adopting COVID-19 response from neighboring countries, result in large variations in mortality rates. A mere three-day delay in the response was estimated to result in more than a doubling in mortality during a single wave.

For any of the six countries, mortality would have differed substantially, had the response of another country been adopted. The order of the resulting cumulative COVID-19 deaths per million, from lowest to the highest, were found for the responses of the Netherlands, Belgium, Denmark, the UK, Germany, and Sweden. This order differs from the observed rates of confirmed COVID-19 deaths, where Denmark and Germany reported the fewest deaths per million before the Netherlands, Sweden, Belgium, and the UK. Actual observed per-capita death rates are determined not only by the response but also by underreporting and the epidemiological situation in the early phase of the pandemic wave: for example, the incidence of infection on 13 March 2020 and the reproduction number before this time varied. In early March 2020, COVID-19 mortality trajectories in the Netherlands, UK, and Belgium slightly outpaced those in Germany, Denmark, and Sweden. This implies, for instance, that a marginally lower incidence of infection in Denmark compared to Netherlands allowed for a slower decline in R_t_, while still resulting in a lower observed mortality rate. Furthermore, large-scale implementation of response measures may lead to the fastest reduction of the reproduction number R_t_ in countries with the highest viral transmission, as in countries with low transmission, the virus may emerge in settings where transmission is harder to control.

Another aspect is that the reproduction number R_t_ without control measures differed between countries, being estimated higher in the Netherlands, Belgium and the UK compared to Germany, Denmark and Sweden. A higher reproduction number R_t_ without control measures requires a more effective response to bring the reproduction number R_t_ below 1. We conducted an additional analysis in which the absolute R_t_ value from other countries was transposed to the Netherlands (instead of the relative reduction of R_t_ without control measures). We found that the order of outcome is not affected (Supplemental Table S1), although the difference between counterfactual and observed number of deaths per million became smaller for responses from Germany, Denmark and Sweden. For instance, with the exchange of the absolute R_t_, the responses of Denmark and the Netherlands resulted in similar mortality instead of a more than two-fold higher mortality with the response of Denmark using the relative reduction in R_t_.

Our finding that small fluctuations in the reproduction number during a fast-growing epidemic can significantly impact mortality rates aligns with previous studies. For example, a UK study estimated a potential 73% decrease in COVID-19 deaths if lockdown measures were implemented one week earlier in spring 2020 [12], while similar analyses for Sweden suggested a 34-40% reduction in deaths by May 2020 with a lockdown similar to Denmark or Norway [13, 14]. Our analysis builds upon the previous study of Mishra et al. [5], incorporating three additional countries: the Netherlands, Belgium and Germany. This expansion extends the intercountry comparisons from 6 to 30, facilitating the comparison of neighboring countries with similar initial trajectory of COVID-19 mortality in the initial wave (Germany similar to Denmark and Sweden, and Netherlands similar to Belgium and the UK). Our finding that the mortality does not only depend on the response of a donor country but also on the characteristics and epidemiological situation of the recipient country was only possible through this larger-scale intercountry comparison.

Our analysis comes with several limitations. Firstly, we applied the same delay between infection and death across countries. While the median delay time from symptom onset to death in the Netherlands was previously estimated at 11 days [15], consistent with the data from England used in this study, this assumption may not hold across all studied countries. Secondly, we relied on exchanging reproduction numbers derived from time series of confirmed COVID-19 deaths, which can be influenced by reporting quality and case definitions. However, alternative national vital statistics data usually report deaths on a weekly basis, while daily data is needed to accommodate the small differences in timing of interventions between the selected countries. Moreover, the reproduction number is a relative measure, implying that the relative comparisons between mortality rates remain unaffected if the level of underreporting is stable over time. The Netherlands had about 35% underreporting of COVID-19 deaths compared to excess deaths in the first wave [15], while Belgium had no underreporting [16]; nonetheless, the Netherlands also peaked earlier than Belgium in hospitalization rates [1], indicating consistent findings across different outcomes.

Careful distinction between the counterfactual assessments and the actual implementation of a different response in another country is essential. Continuous epidemic monitoring will lead to intensified control measures when the current set proves insufficient to curb rising mortality, and to relaxation of measures when mortality is low or control appears overly stringent. However, inherent to delays in monitoring the impact of these measures, from implementation to decreasing rates of hospitalization or mortality due to COVID-19, we believe that adaptive control likely will not alter the presented outcomes. Additionally, compliance to responses also varies between countries, possibly influenced by differences in public trust in governmental institutions. Public support for measures is also influenced by response strategies in neighboring countries or the potential risk of exceeding health care capacities, further underscore the complexity.

These findings should also be interpreted considering the limited duration of the study period and available knowledge at the time. For instance, during the first COVID-19 wave, it was unknown that an effective vaccine would become available within a year, that individuals with mild infections could suffer from post-COVID conditions, and that several new, more transmissible variants would emerge within two years, each with different illness severity. Moreover, different responses affect the speed of (herd) immunity buildup, potentially leading to varied outcomes when evaluating the same strategies over a longer period.

Our study contributes to discussions about the merits of the different approaches taken in European countries. They demonstrate that the outcome of response is determined not only by the response itself but also to a large extend by small differences in the initial epidemiological situation in each country. Some countries had relatively low mortality rates for any of the six responses evaluated here, and these countries could afford a response that was less stringent; other countries faced relatively high mortality rates for any of the six response evaluated here, and these countries could ill afford a less stringent response. This underscores that a proper response has to be carefully tailored to the epidemiological situation in each country.

## Conclusion

This analysis shows that in a fast-growing epidemic, small differences in the timing and effectiveness of measures can result in large variations in mortality. For most countries, adopting a response from a neighboring European country during the first COVID-19 pandemic wave in 2020 resulted in an outcome that differed greatly from the outcome observed in the neighboring country. The responses from the six countries studied here revealed a seven-fold to twelve-fold difference between lowest and highest mortality rate. Differences in country characteristics and initial epidemiological situations cause that the outcome of the response in a particular country does not necessarily result in mortality as in that other country; a response must always be tailor-made. A three-day delay of the response was estimated to more than double mortality. These findings provide useful insights in the evaluation of COVID-19 responses and for strategic planning on how to minimize the disease burden of future pandemics.

## Statements

### Acknowledgements

We thank Matthias an der Heiden of the Robert Koch Institute, Germany, for providing the time series of COVID-19 deaths in Germany.

### Authors contributions

The project was conceptualized by PTdB, FM, and JW. Data collection was performed by PTdB and GL. PTdB and GL adapted the existing model code for the current analysis, which was critically reviewed by FM. The initial draft of the manuscript was written by PTdb, which was critically revised by FM, GL and JW. All authors have read and approved of the final manuscript.

### Ethical statement

Ethical approval was not required for this study as all data used within this work was part of routine surveillance

### Data availability

Timeseries of deaths by date of death will become available online

## Supplementary materials

Supplementary materials belonging to the manuscript:

Evaluating the COVID-19 responses of Belgium, Denmark, Germany, the Netherlands, Sweden and the United Kingdom, February-June 2020: A counterfactual modelling study

---

**Figure S1:**
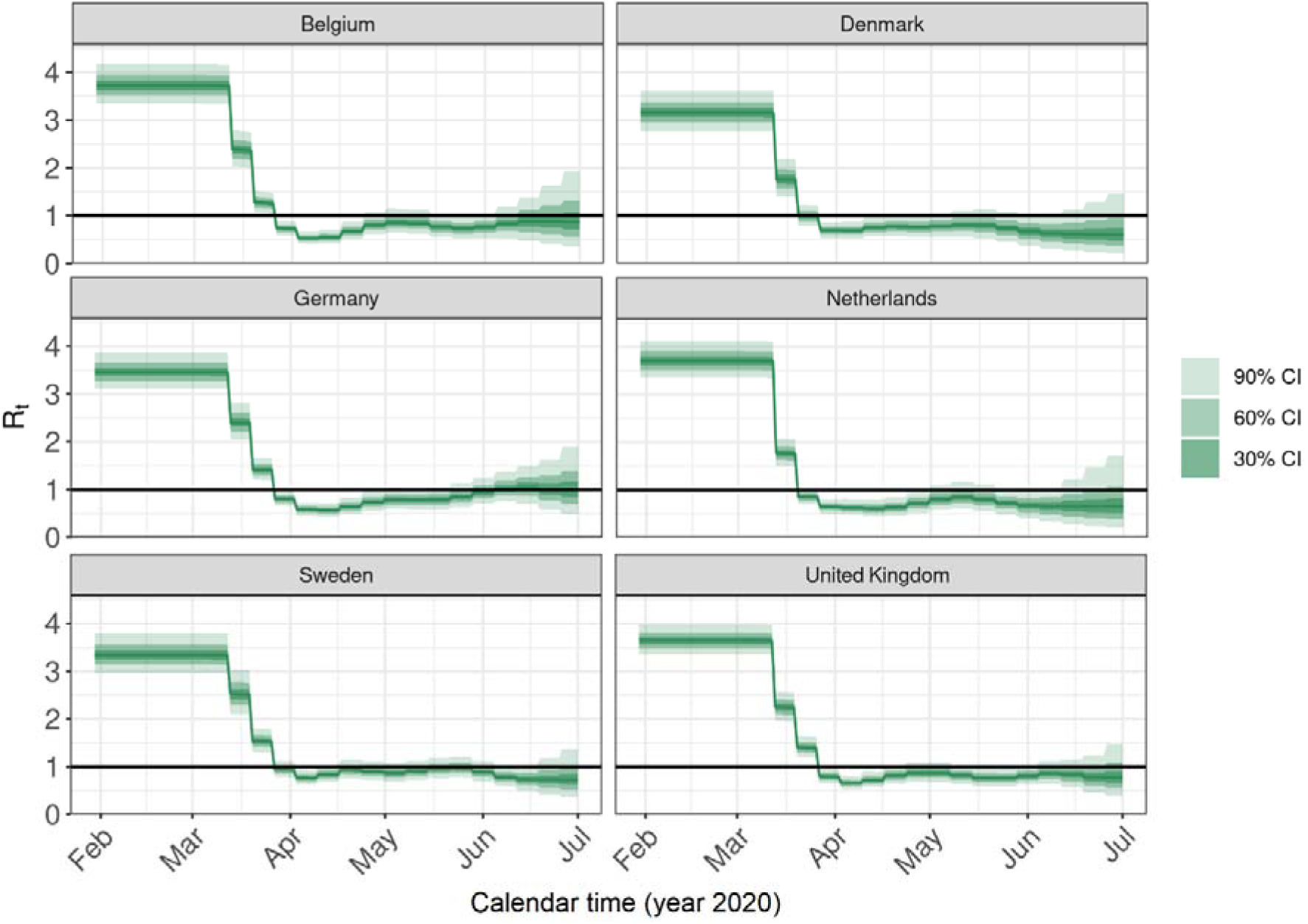
Estimated time-varying reproduction number (R_t_) per country in the period February, 2020, up to and including June, 2020, with their 30%, 60% and 90% credible intervals. The reproduction numbers are estimated using daily numbers of confirmed deaths by date of death. The R_t_ estimates for Denmark, Sweden and the UK deviate from the analysis of Mishra et al. [5]. This is due to the use of a different generation time.

**Figure S2:**
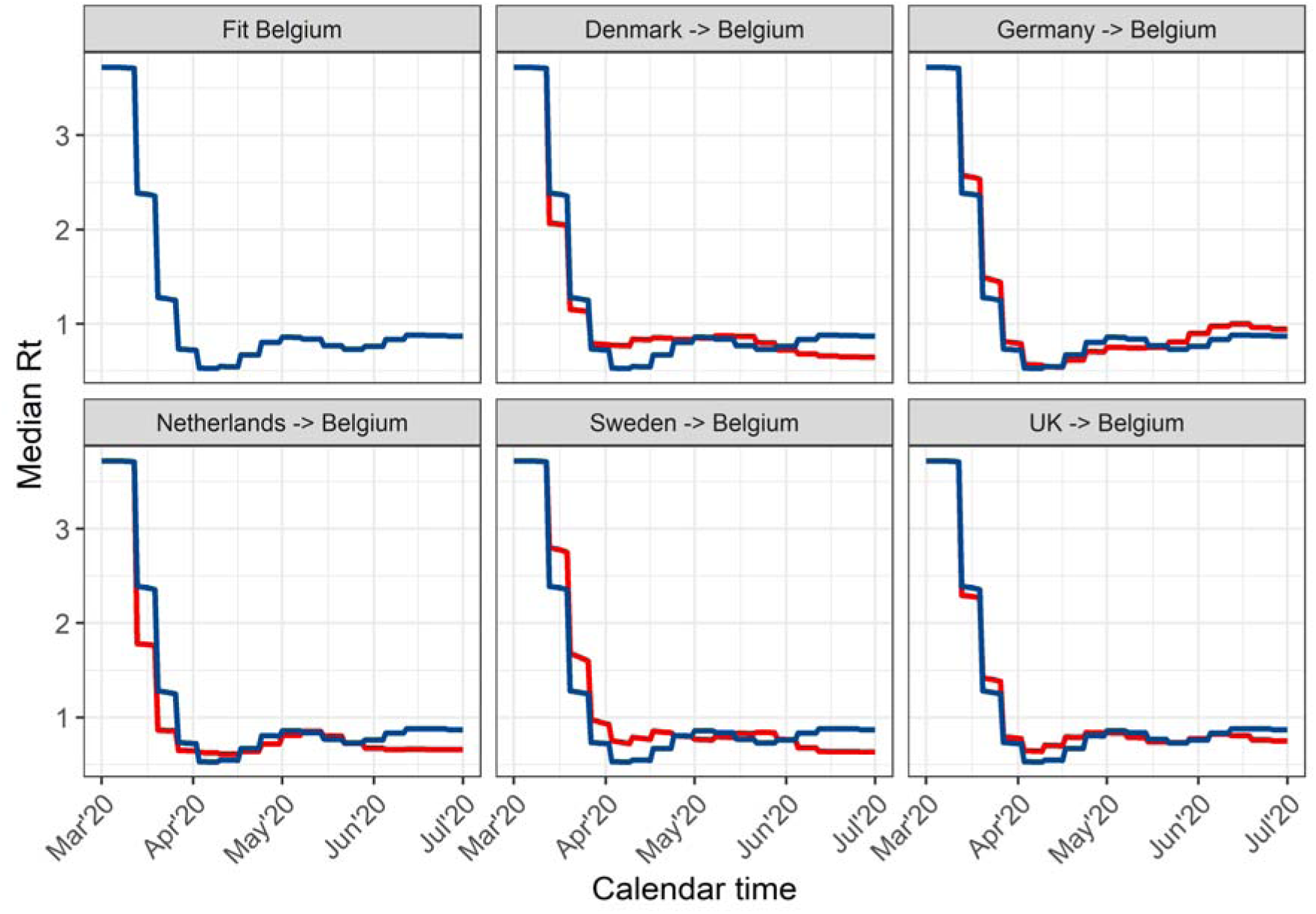
The estimated median reproduction number R_t_ for the Belgium using mortality data (blue lines), and the R_t_ for the different counterfactual analyses (red lines), after transferring the relative reduction in reproduction number from Denmark, Germany, the Netherlands, Sweden, and the United Kingdom (UK) to Belgium between March 13, 2020, and July 1, 2020.

**Figure S3:**
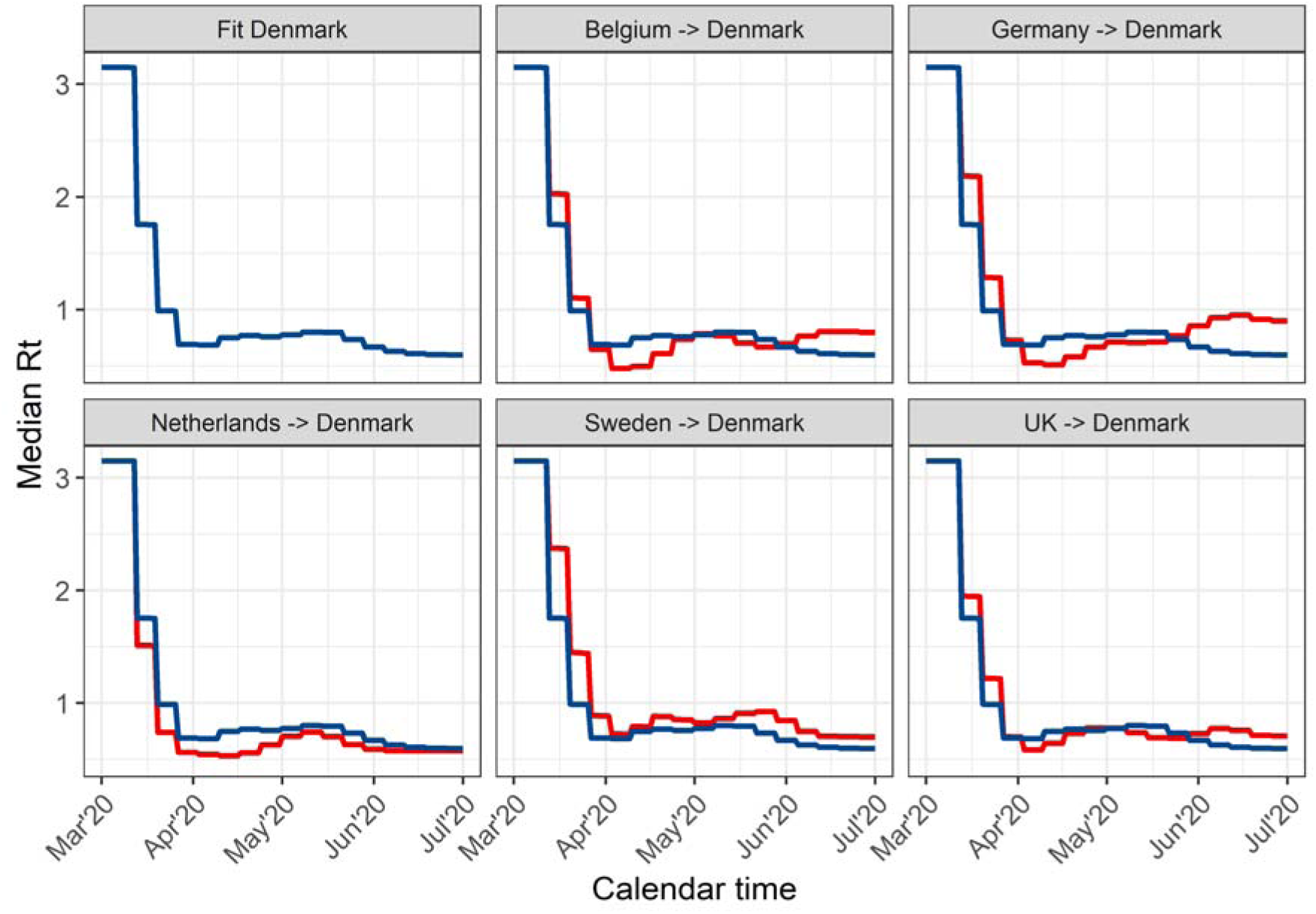
The estimated median reproduction number R_t_ for Denmark using mortality data (blue lines), and the R_t_ for the different counterfactual analyses (red lines), after transferring the relative reduction in reproduction number from Belgium, Germany, the Netherlands, Sweden, and the United Kingdom (UK) to the Denmark between March 13, 2020, and July 1, 2020.

**Figure S4:**
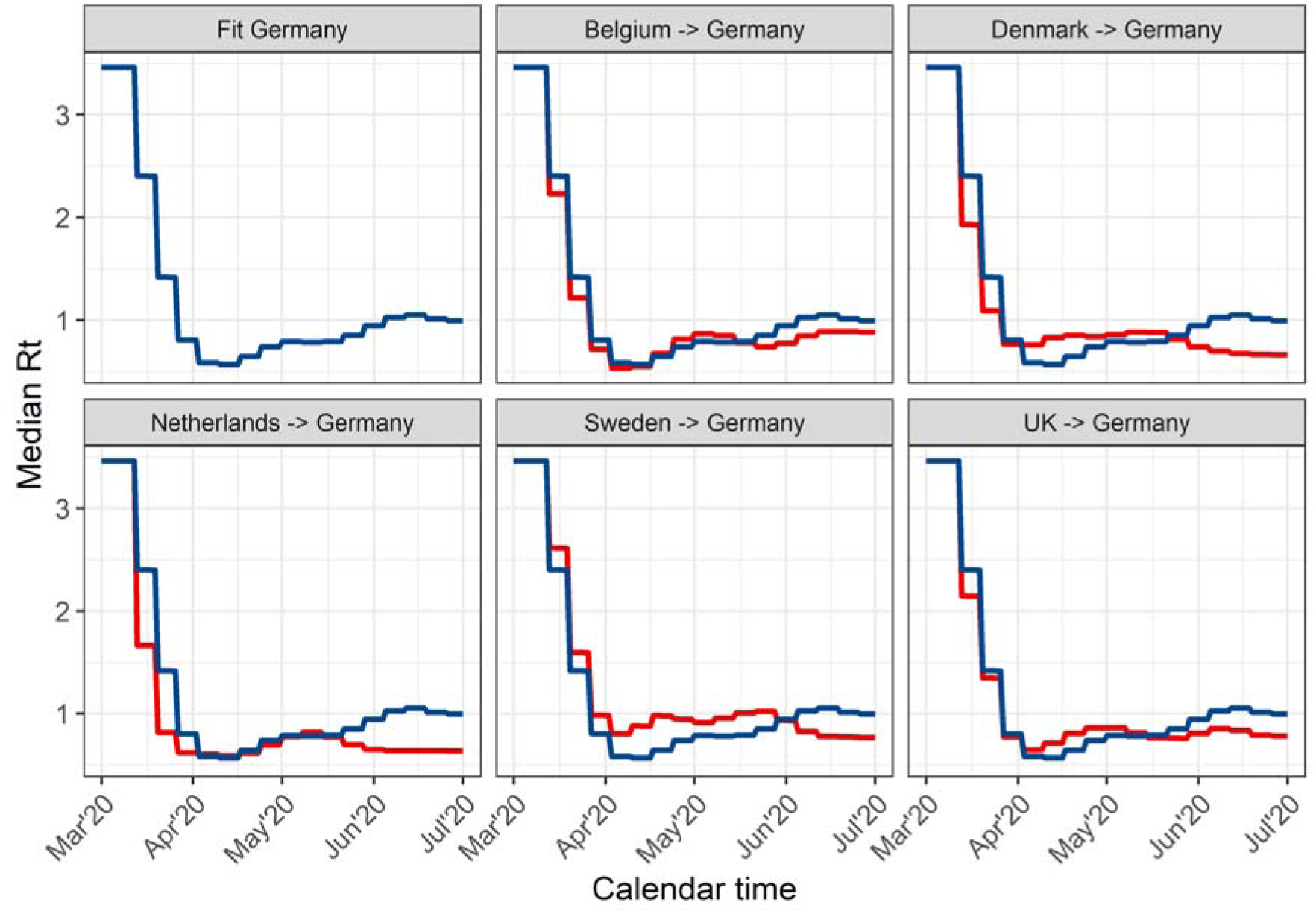
The estimated median reproduction number R_t_ for Germany using mortality data (blue lines), and the R_t_ for the different counterfactual analyses (red lines), after transferring the relative reduction in reproduction number from Belgium, Denmark, Netherlands, Sweden, and the United Kingdom (UK) to Germany between March 13, 2020, and July 1, 2020.

**Figure S5:**
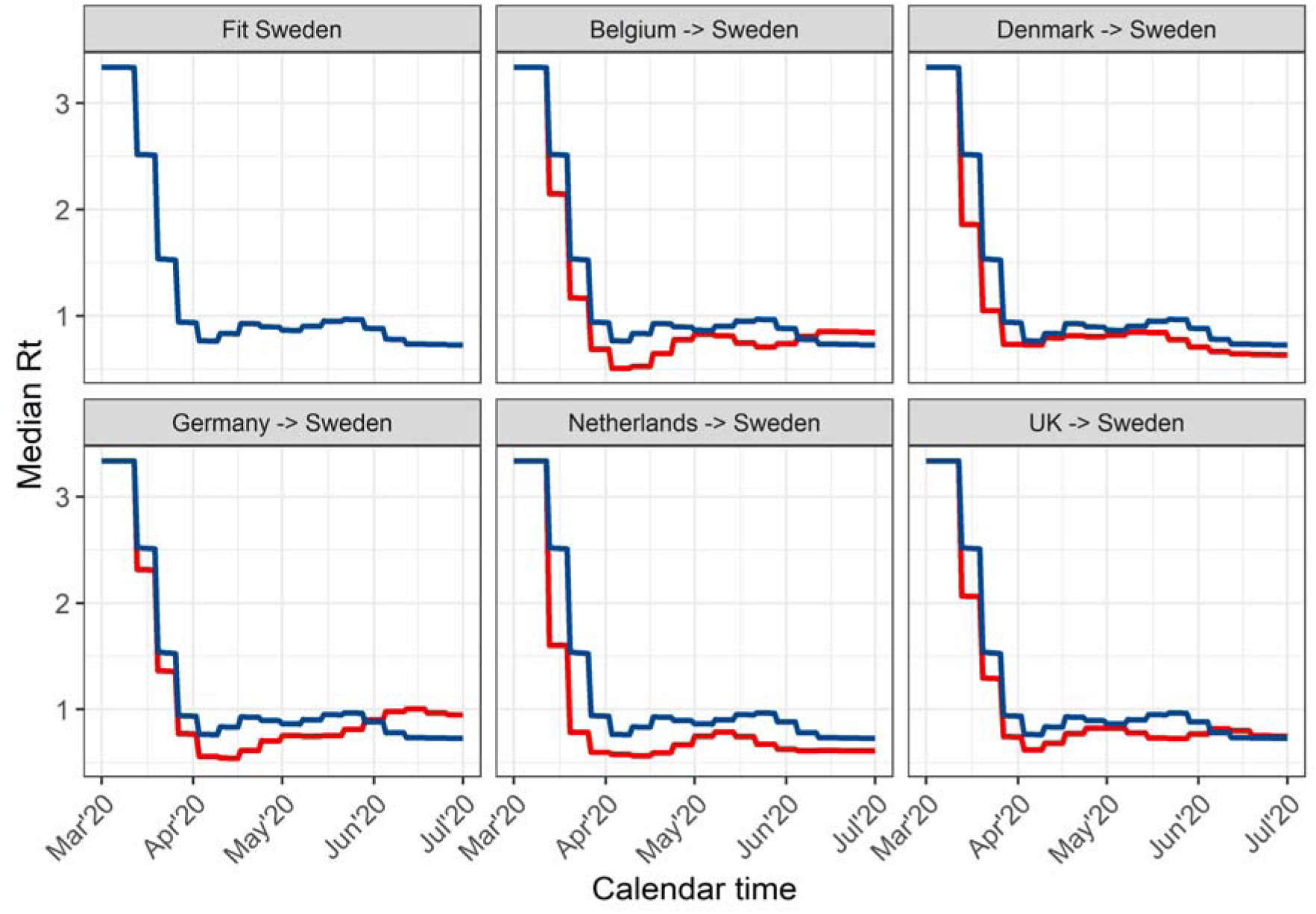
The estimated median reproduction number R_t_ for Sweden using mortality data (blue lines), and the R_t_ for the different counterfactual analyses (red lines), after transferring the relative reduction in reproduction number from Belgium, Denmark, Germany, the Netherlands, and the United Kingdom (UK) to Sweden between March 13, 2020, and July 1, 2020.

**Figure S6:**
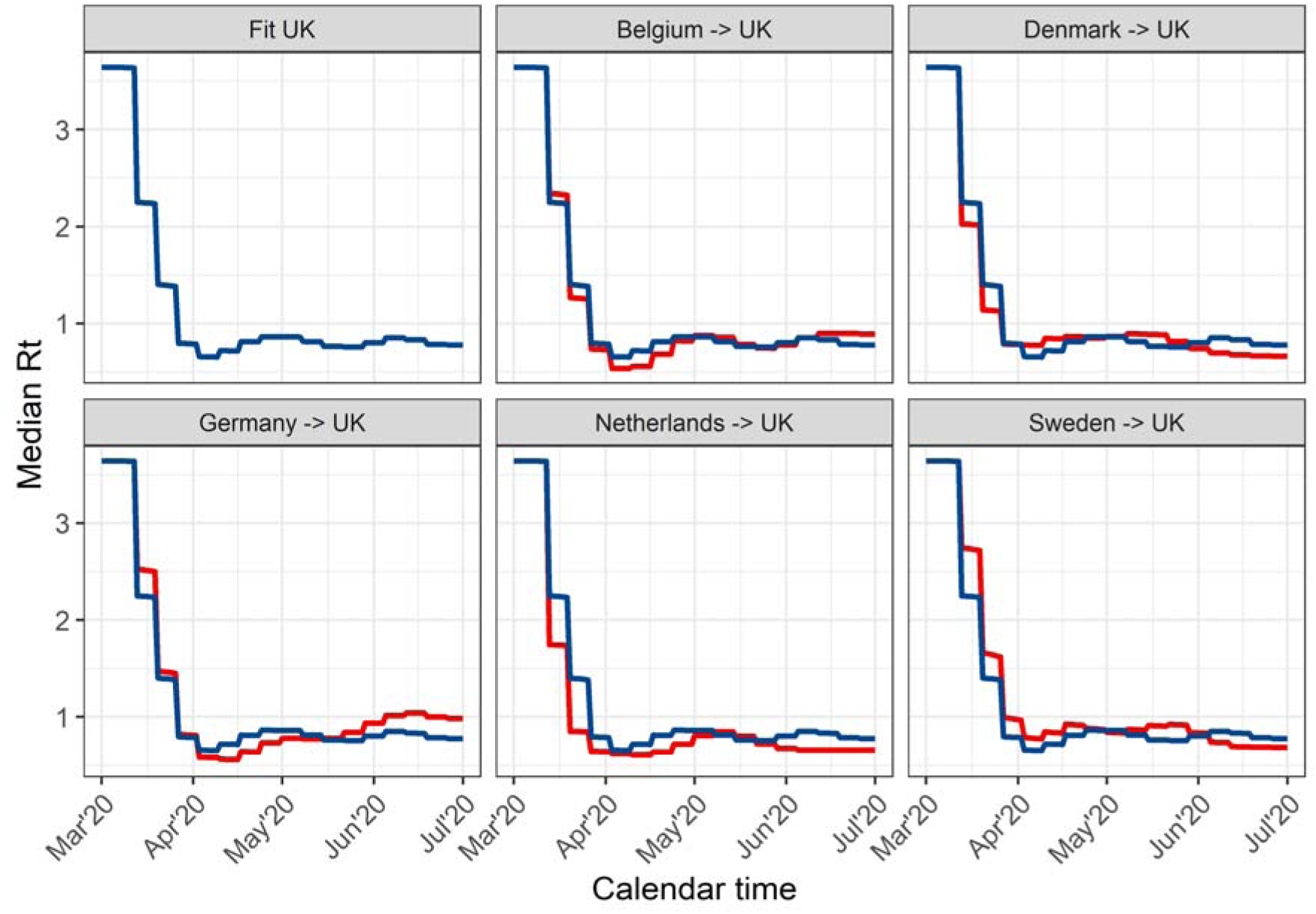
The estimated median reproduction number R_t_ for the United Kingdom (UK) using mortality data (blue lines), and the R_t_ for the different counterfactual analyses (red lines), after transferring the relative reduction in reproduction number from Belgium, Denmark, Germany, the Netherlands, and Sweden, to the UK between March 13, 2020, and July 1, 2020.

**Figure S7:**
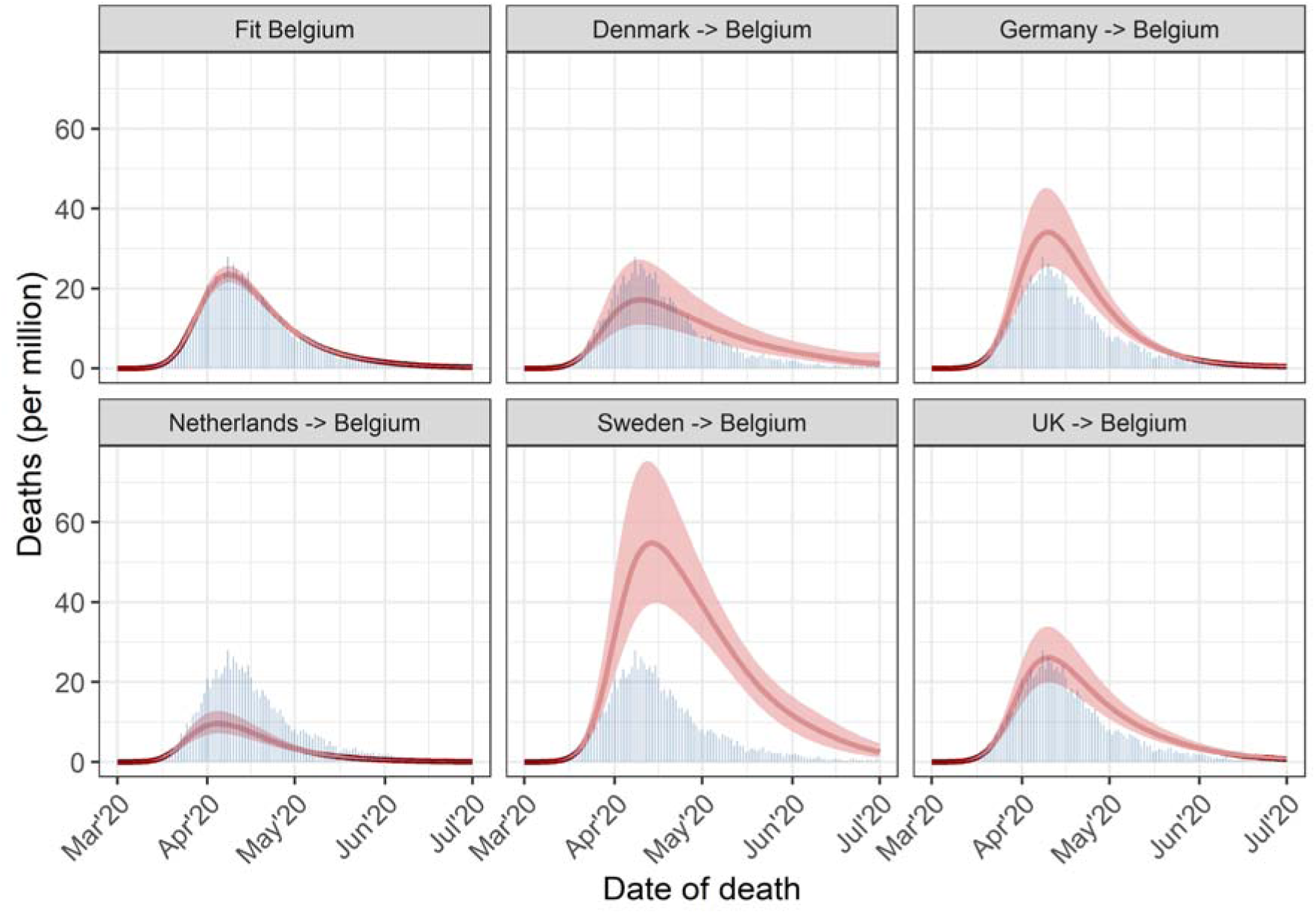
Estimated median number of daily deaths with 95% credible intervals for Belgium, showing the fit to observed data (blue bars) and the counterfactual analyses, involving the transfer of the relative reduction in reproduction number from Denmark, Germany, the Netherlands, Sweden, and the United Kingdom (UK) to Belgium in the period March 13, 2020, to July 1, 2020.

**Figure S8:**
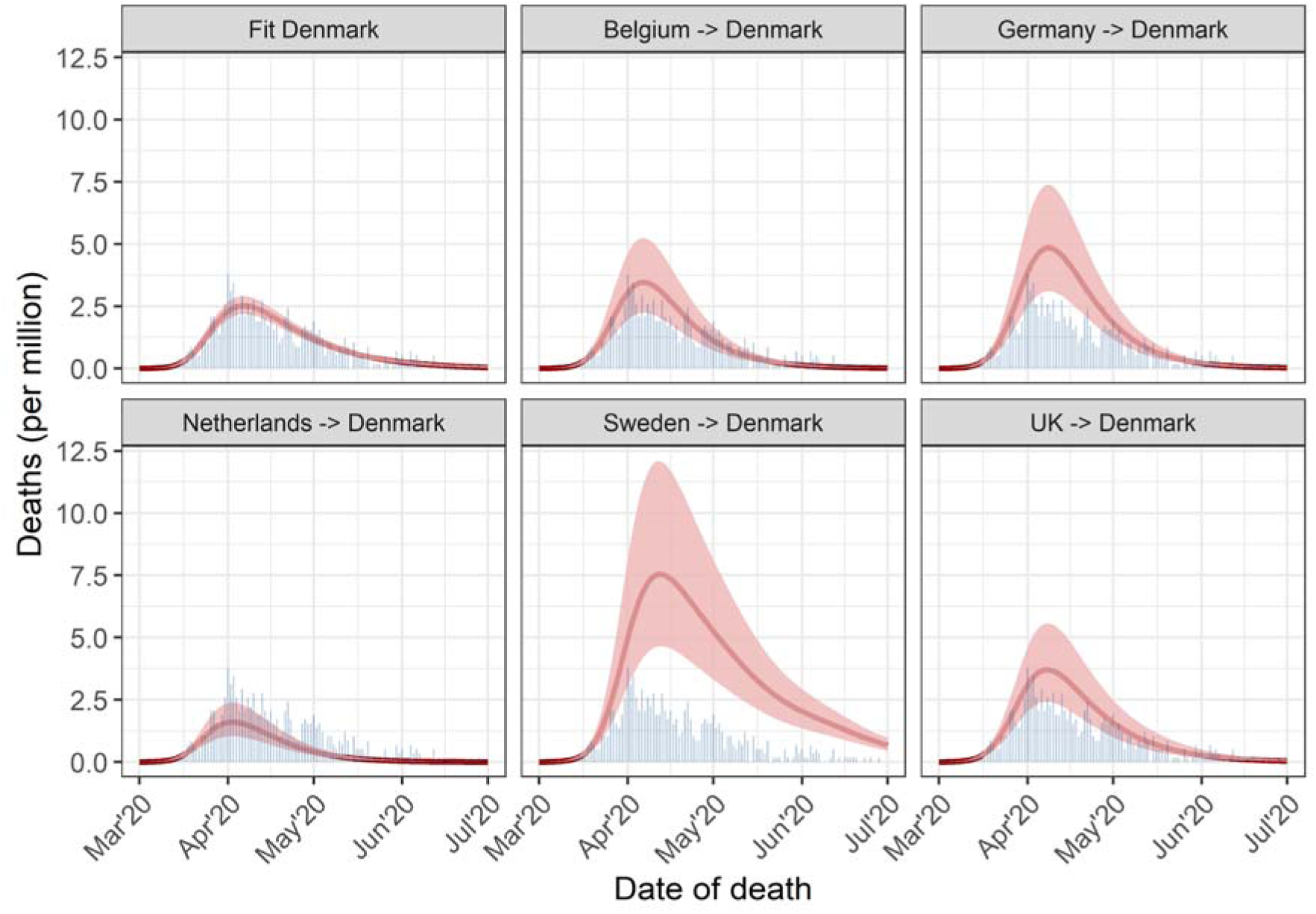
Estimated median number of daily deaths with 95% credible intervals for Denmark, showing the fit to observed data (blue bars) and the counterfactual analyses, involving the transfer of the relative reduction in reproduction number from Belgium, Germany, the Netherlands, Sweden, and the United Kingdom (UK) to Denmark in the period March 13, 2020, to July 1, 2020.

**Figure S9:**
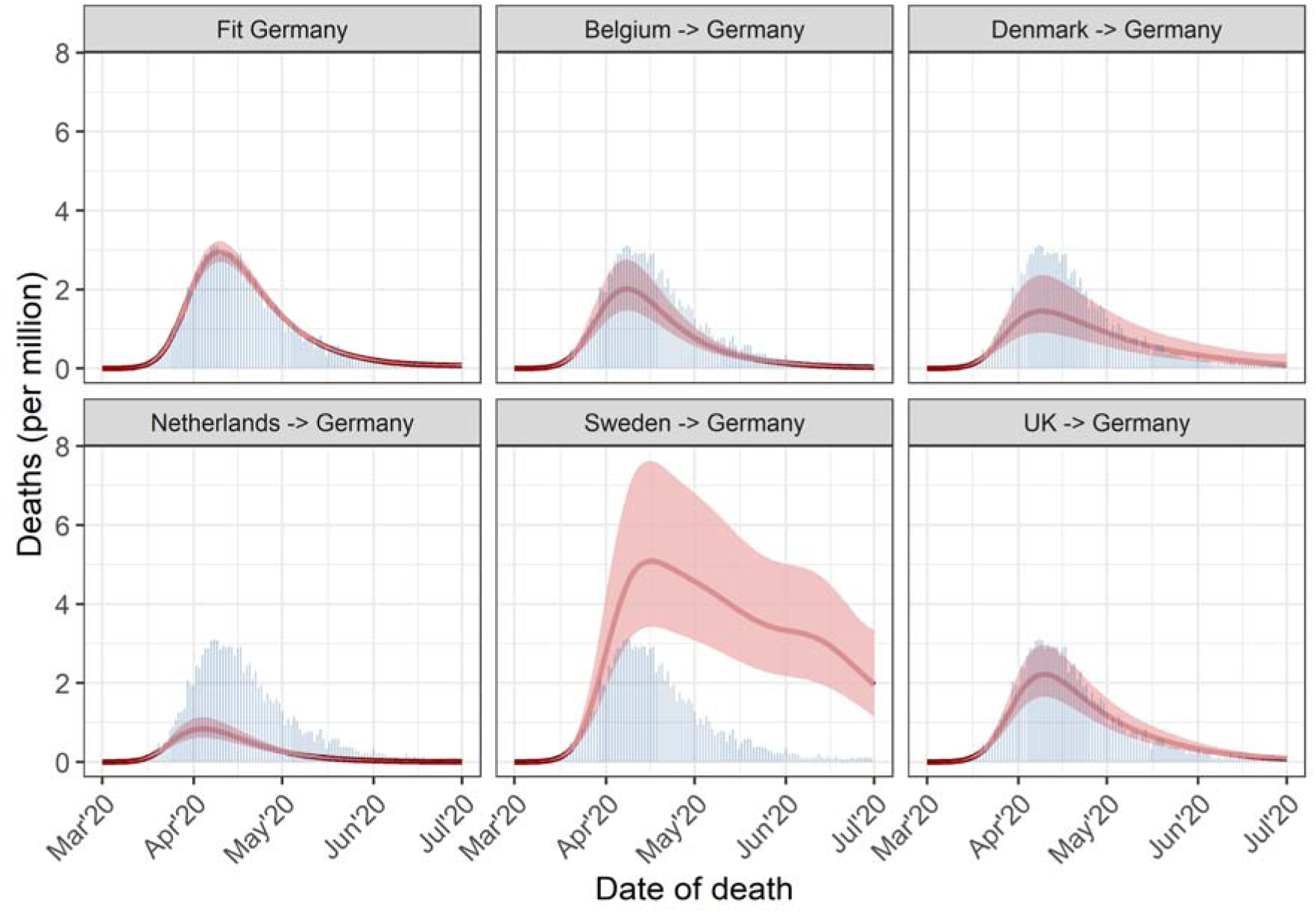
Estimated median number of daily deaths with 95% credible intervals for Germany, showing the fit to observed data (blue bars) and the counterfactual analyses, involving the transfer of the relative reduction in reproduction number from Belgium, Denmark, the Netherlands, Sweden, and the United Kingdom (UK) to Germany in the period March 13, 2020, to July 1, 2020.

**Figure S10:**
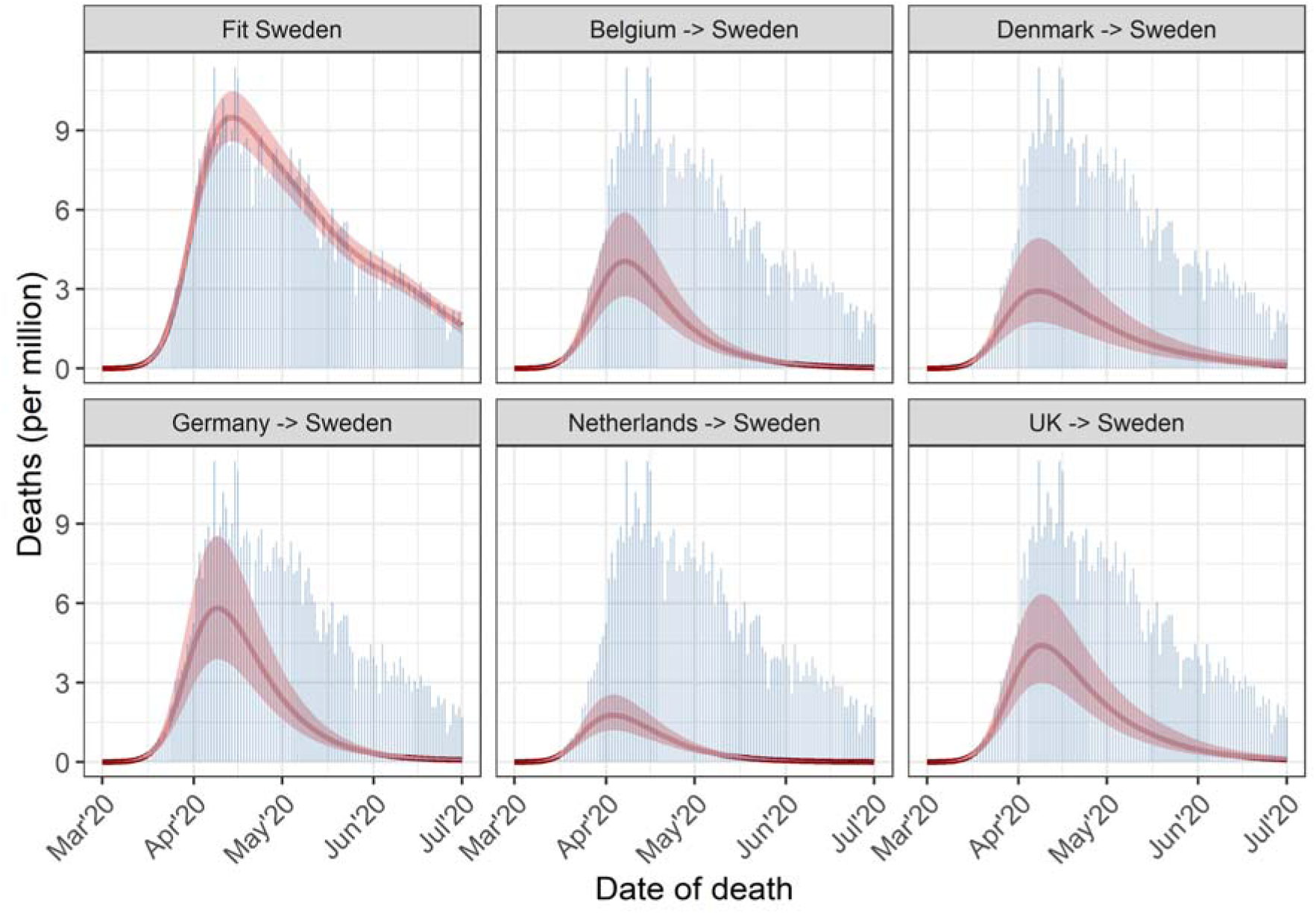
Estimated median number of daily deaths with 95% credible intervals for Sweden, showing the fit to observed data (blue bars) and the counterfactual analyses, involving the transfer of the relative reduction in reproduction number from Belgium, Denmark, Germany, the Netherlands, and the United Kingdom (UK) to Sweden in the period March 13, 2020, to July 1, 2020.

**Figure S11:**
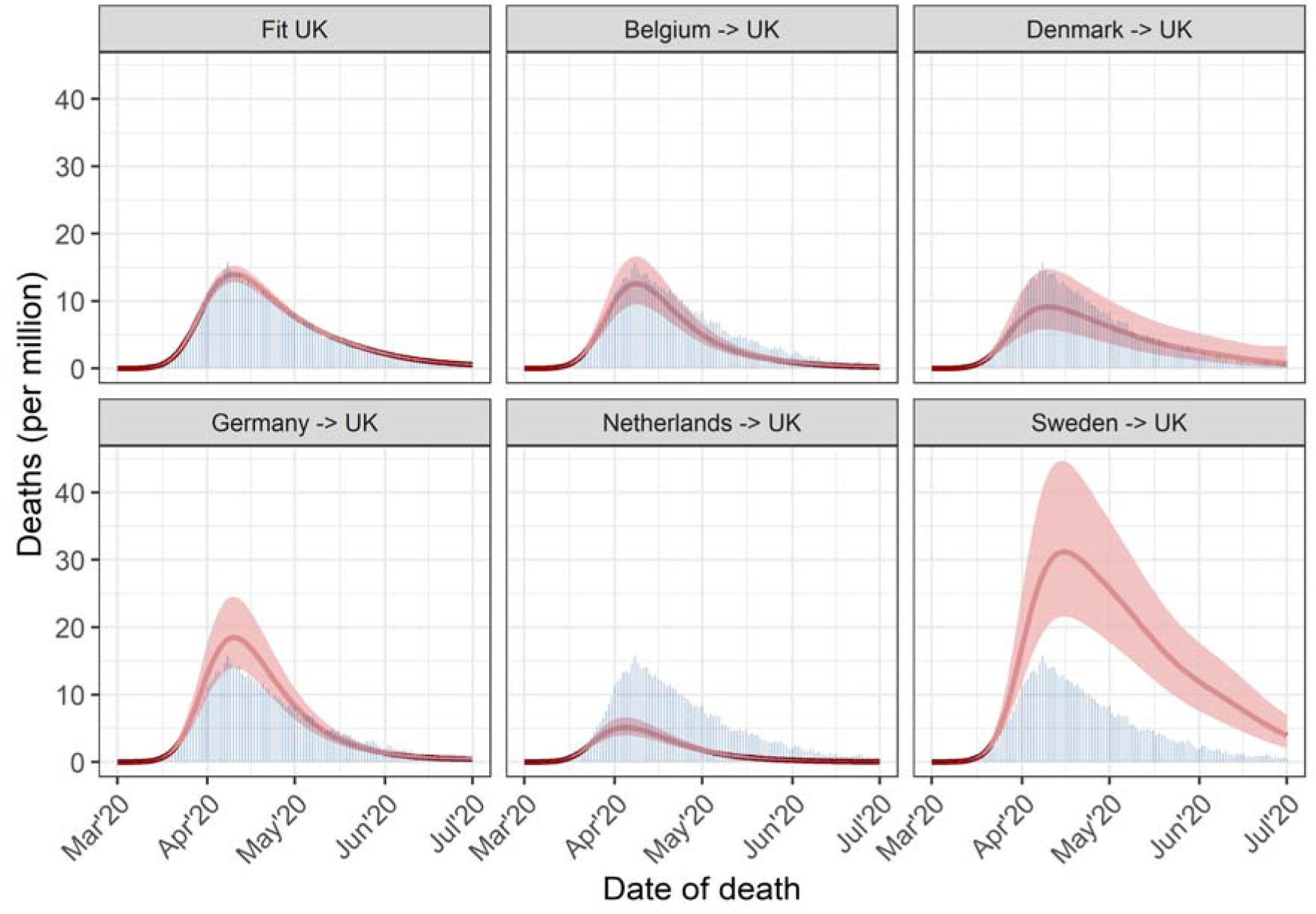
Estimated median number of daily deaths with 95% credible intervals for the United Kingdom (UK), showing the fit to observed data (blue bars) and the counterfactual analyses, involving the transfer of the relative reduction in reproduction number from Belgium, Denmark, Germany, the Netherlands, and Sweden, to the UK in the period March 13, 2020, to July 1, 2020.

**Figure S12:**
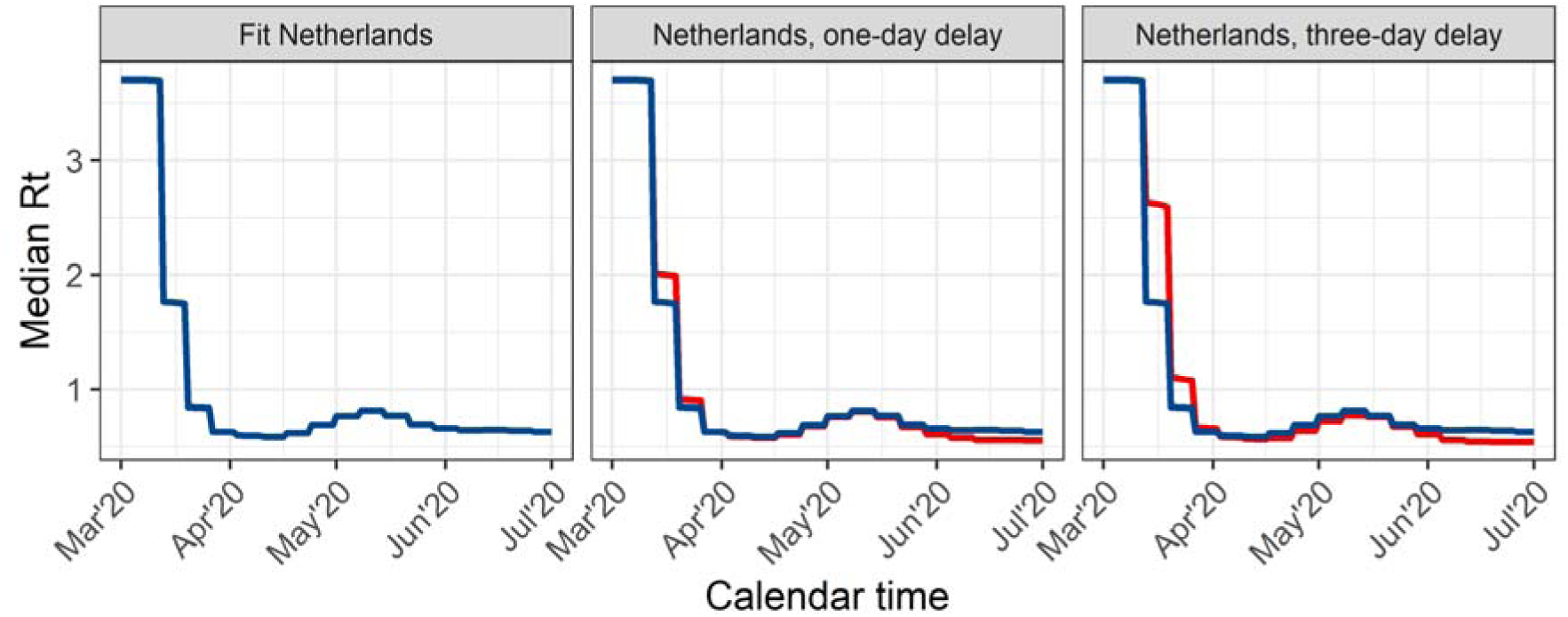
Estimated median reproduction number R_t_ for the Netherlands using mortality data (blue lines), and the R_t_ for the different counterfactual analyses (red lines), in the period March 13, 2020, to July 1, 2020, if the response measures were taken one day later or three days later.

**Table S1:** Scenario analysis of cumulative COVID-19-attributed deaths per million inhabitants per country until 1^st^ of July 2020. Diagonal elements show observed mortality, off-diagonal elements show mortality (median with 95% credible intervals) for counterfactual strategies.. In this approach, the exact absolute values of the reproduction number R_t_ from the ‘donor’ country are transposed to the ‘recipient’ country, starting on 13^th^ March, 2020.

| Response as implemented in a country (donor country) | Outcome of response when applied to a country (recipient country) |  |  |  |  |  |  |  |  |  |  |  |
| --- | --- | --- | --- | --- | --- | --- | --- | --- | --- | --- | --- | --- |
|  | Belgium |  | Denmark |  | Germany |  | Netherlands |  | Sweden |  | United Kingdom |  |
|  | Cum. deaths per million | MF<sup>a</sup> | Cum. deaths per million | MF<sup>a</sup> | Cum. deaths per million | MF<sup>a</sup> | Cum. deaths per million | MF<sup>a</sup> | Cum. deaths per million | MF<sup>a</sup> | Cum. deaths per million | MF<sup>a</sup> |
| **Belgium** | 840 | 2.4 | 223 [145-332] | 2.6 | 99 [72-134] | 2.7 | 861 [671-1101] | 2.4 | 219 [148-316] | 2.6 | 497 [389-637] | 2.5 |
| **Denmark** | 418 [286-620] | 1.2 | 105 | 1.2 | 46 [30-70] | 1.2 | 431 [300-632] | 1.2 | 103 [64-166] | 1.5 | 240 [168-354] | 1.2 |
| **Germany** | 920 [700-1201] | 2.6 | 244 [158-364] | 2.8 | 108 | 2.9 | 947 [734-1219] | 2.6 | 240 [162-348] | 2.8 | 546 [424-704] | 2.7 |
| **Netherlands** | 350 [267-457] | 1 | 86 [56-128] | 1 | 37 [28-50] | 1 | 361 | 1 | 85 [57-121] | 1 | 199 [158-253] | 1 |
| **Sweden** | 1609 [1238-2095] | 4.6 | 555 [362-822] | 6.5 | 266 [186-384] | 7.2 | 1644 [1281-2121] | 4.6 | 546 | 6.4 | 1080 [829-1417] | 5.4 |
| **UK** | 978 [781-1213] | 2.8 | 276 [185-396] | 3.2 | 124 [94-162] | 3.4 | 1005 [817-1228] | 2.8 | 272 [190-376] | 3.2 | 599 | 3.0 |

<sup>a</sup>: MF is the multiplicative factor in deaths compared to the number of deaths estimated with the response of the Netherlands (= 1). The counterfactual comparisons between Denmark, Sweden and the UK slightly deviate from the analysis of Mishra et al. [5] due to the use of a different generation time.

## References

1. Mathieu E, Ritchie H, Rodés-Guirao L, Appel C, Giattino C, Hasell J, Macdonald B, Dattani S, Beltekian D, Ortiz-Ospina E, Roser M. Coronavirus Pandemic (COVID-19) 2020 https://ourworldindata.org/coronavirus. Accessed at 8 December 2023.

2. Brauner JM, Mindermann S, Sharma M, Johnston D, Salvatier J, Gavenciak T, Stephenson AB, Leech G, Altman G, Mikulik V et al. Inferring the effectiveness of government interventions against COVID-19. Science 2021, 371(6531).

3. Sharma M, Mindermann S, Rogers-Smith C, Leech G, Snodin B, Ahuja J, Sandbrink JB, Monrad JT, Altman G, Dhaliwal G et al. Understanding the effectiveness of government interventions against the resurgence of COVID-19 in Europe. Nat Commun 2021, 12(1):5820.

4. Hale T, Angrist N, Goldszmidt R, Kira B, Petherick A, Phillips T, Webster S, Cameron-Blake E, Hallas L, Majumdar S, Tatlow H. A global panel database of pandemic policies (Oxford COVID-19 Government Response Tracker). Nat Hum Behav 2021, 5(4):529–538.

5. Mishra S, Scott JA, Laydon DJ, Flaxman S, Gandy A, Mellan TA, Unwin HJT, Vollmer M, Coupland H, Ratmann O et al. Comparing the responses of the UK, Sweden and Denmark to COVID-19 using counterfactual modelling. Sci Rep 2021, 11(1):16342.

6. Lison A, Banholzer N, Sharma M, Mindermann S, Unwin HJT, Mishra S, Stadler T, Bhatt S, Ferguson NM, Brauner J, Vach W. Effectiveness assessment of non-pharmaceutical interventions: lessons learned from the COVID-19 pandemic. Lancet Public Health 2023, 8(4):e311–e317.

7. Sciensano Epistat. COVID-19 2022 https://epistat.sciensano.be/covid/. Accessed at 24 May 2022.

8. Backer JA, Eggink D, Andeweg SP, Veldhuijzen IK, van Maarseveen N, Vermaas K, Vlaemynck B, Schepers R, van den Hof S, Reusken CB, Wallinga J. Shorter serial intervals in SARS-CoV-2 cases with Omicron BA.1 variant compared with Delta variant, the Netherlands, 13 to 26 December 2021. Euro Surveill 2022, 27(6).

9. Lauer SA, Grantz KH, Bi Q, Jones FK, Zheng Q, Meredith HR, Azman AS, Reich NG, Lessler J. The Incubation Period of Coronavirus Disease 2019 (COVID-19) From Publicly Reported Confirmed Cases: Estimation and Application. Ann Intern Med 2020, 172(9):577–582.

10. Fraser C. Estimating individual and household reproduction numbers in an emerging epidemic. PLoS One 2007, 2(8):e758.

11. Wallinga J, Lipsitch M. How generation intervals shape the relationship between growth rates and reproductive numbers. Proc Biol Sci 2007, 274(1609):599-604.

12. Arnold KF, Gilthorpe MS, Alwan NA, Heppenstall AJ, Tomova GD, McKee M, Tennant PWG. Estimating the effects of lockdown timing on COVID-19 cases and deaths in England: A counterfactual modelling study. PLoS One 2022, 17(4):e0263432.

13. Born B, Dietrich AM, Muller GJ. The lockdown effect: A counterfactual for Sweden. PLoS One 2021, 16(4):e0249732.

14. Latour C, Peracchi F, Spagnolo G. Assessing alternative indicators for Covid-19 policy evaluation, with a counterfactual for Sweden. PLoS One 2022, 17(3):e0264769.

15. de Boer PT, van de Kassteele J, Vos ERA, van Asten L, Dongelmans DA, van Gageldonk- Lafeber AB, den Hartog G, Hofhuis A, van der Klis F, de Lange DW et al. Age-specific severity of severe acute respiratory syndrome coronavirus 2 in February 2020 to June 2021 in the Netherlands. Influenza Other Respir Viruses 2023, 17(8):e13174.

16. Molenberghs G, Faes C, Verbeeck J, Deboosere P, Abrams S, Willem L, Aerts J, Theeten H, Devleesschauwer B, Bustos Sierra N et al. COVID-19 mortality, excess mortality, deaths per million and infection fatality ratio, Belgium, 9 March 2020 to 28 June 2020. Euro Surveill 2022, 27(7).

